# A study of sociodemographic and clinical profiles of HIV-1 infected North Indian patients

**DOI:** 10.1101/2021.03.05.21252925

**Authors:** Sushanta Kumar Barik, Srikanth Prasad Tripathy, Ramesh Karunaianantham, Sathyamurthi Pattabiraman, Virendra Singh Yadav, Tej Pal Singh, Rekha Tandon, Sheetal Tomar, Srikanta Jena, Shripad A Patil, Keshar Kunja Mohanty

**Affiliations:** ICMR- National JALMA Institute for Leprosy and Other Mycobacterial Diseases, Agra, Uttar-Pradesh-282004, India.; ICMR-National Institute for Research in Tuberculosis, Chetpet-600031, Chennai, Tamil-Nadu, India.; Sarojini Naidu Medical College, Agra-282003, Uttar-Pradesh, India.; Ravenshaw University, Cuttack-753003, Odisha, India.

**Keywords:** HIV, AIDS, ART, Viral load, CD4

## Abstract

The aim of this study was to analyze the sociodemographic, clinical, immunologic, and risk factors of drug-resistant (>1000 copies/ml) and virologically suppressed (<1000 to <40 copies /ml to target not detected levels) patients at the ART center, Sarojini Naidu Medical College, Agra, North India. A total of 193 patients on first-line antiretroviral therapy were included those were recruited from December 2009 to November 2016. The patients included in this study had a CD4 cell count of ≤ 350 cubic/mm. The details of Demographic, clinical, and social factors were collected in a patient information leaflet and statistically analyzed. After viral load, two groups of patients were observed. The drug-resistant group (N=58) had a viral load ≥1000 copies/ml and virologically suppressed group (N=135) had a viral load <1000 copies/ml to target not detected level. A comparison statement result was presented in both groups of drug-resistant and virologically suppressed patients. Males were observed in the highest frequency (42, 72.41%). The heterosexual mode of transmission was predominant (40, 68.96%, and 98,72.59%). The highest number of married couples and illiterates in the two groups. Tuberculosis was observed in the highest numbers in the two groups. The analysis of socioeconomic factors of North Indian patients will be a concern issue in the demographic profiles of HIV-1 patients. The clinical features, analysis would help clinicians in further therapy implementation, monitoring CD4 and Viral load counts of North Indian patients.

## Introduction

The human immunodeficiency virus (HIV) was transmitted among all types of rational groups of people, but mostly effective in weaker sections of the people in the society [1]. Socio-demographic and economic status is associated with the country’s burden of disease. This socio-demographic and clinical features often gives information on the flow rate of disease in a region and statistically evaluates the depth of the disease in a population [2]. The clinical and socio-demographic profile of patients attending the anti-retroviral therapy (ART) center would be helpful in acquired immunodeficiency syndrome (AIDS) management. Economic status and productive age groups with the mode of transmission are essential during a tertiary care hospital study [3]. Acquired immunodeficiency syndrome is a life-threatening illness with depression and anxiety. Socioeconomic burden with co-morbidity symptoms is the cause of depression and anxiety in AIDS patients [4]. HIV-1 infection and disease progression are regulated by host and viral factors. The documentation of socio-demographic and epidemiologic features could be useful for improvement of current HIV-1 prevention, monitoring, and therapeutic programs in a particular region [5]. Most of the HIV-infected population is from lower socioeconomic classes, as labors and reproductive age groups are the major risk factors for HIV transmission [6]. Several clinical and epidemiological studies have suggested that HIV transmission in low viral load copies <1000/ml. Therefore, the analysis of the socio-demographic, clinical features, and risk factors are very essential to know about the transmission of HIV patients in a particular region.

The use of a fixed-dose combination of first-line anti-retroviral therapy is a critical tool in terms of safety and efficacy during highly active antiretroviral treatment in a resource-limited setting [7]. Different sampling methods would essential in the HIV surveillance program in a resource-poor setting [8]. Anti-retroviral therapy treatment and monitoring, and patient characteristics have been reported in Sub-Saharan Africa, Asia, and Latin America in resource-limited settings [9].

Therefore, we attempted to analyze the socio-demographic, economic factors, risk factors, and clinical profiles of drug-resistant (DR) and virologically suppressed (VS) first-line ART patients in the ART center, Sarojini Naidu Medical College, Agra, North India. These observations give the cause of the impact of sociodemographic factors, socioeconomic factors, risk factors, and clinical factors influencing HIV transmission in North Indian patients

## Materials and Methods

### Study participants

At the Antiretroviral Therapy Center, Sarojini Naidu Medical College, Agra, North India, 199 patients were recruited on first-line antiretroviral therapy from December 2009 to November 2016. A total of 193 patients whose CD4 cell count was around 350 in each six months during the treatment of first-line antiretroviral therapy.

### Ethics approval

The study protocol was approved by the institutional ethics committee. The Institute ethics committee is constituted according to the guidelines directed by the Indian Council of Medical Research [10].

### Data Collection

The questionnaire was filled by physicians/counselors at the time of enrollment. All socio-demographic features like age, sex, weight, marital status, education, job, monthly income, risk factors, habit of alcohol, habit of smoking, habit of tobacco, clinical profiles like CD4 counts, anti-retroviral therapy, adherence, WHO stages, opportunistic infections, tuberculosis treatment, condom use, pregnancy, hepatitis-B virus, and hepatitis-C virus infection of first-line antiretroviral therapy patients were noted in a patient information leaflet formulated by Sushanta et al. 2018 [11].

### Sample Collection

A 10ml blood sample was collected from the first-line ART patients whose CD4 counts oscillated <350 in the ART center, Sarojini Naidu Medical College, Agra. CD4 cells (FACS count, Becton Dickinson, Mount View, CA) count was performed after enrollment. CD4 counts were performed every six months thereafter during the month of visit for ART in this ART center, Sarojini Naidu Medical College, Agra. Blood samples were shifted in a cold chain setup to the immunology laboratory at the National JALMA Institute for Leprosy and Other Mycobacterial Diseases, Taj-Ganj, Agra, Uttar-Pradesh. Plasma was isolated at 2000 rpm for 10 min at 4°C and stored at −80°C. The plasma samples were then shifted under a well setup cold chain condition to the HIV/AIDS laboratory at the National Institute for Research in Tuberculosis, Chennai for viral load and genotyping.

### Viral load

Viral load (VL) was done by using the Abbott automated m2000rt instrument. Low viral load copies (<1000/ml) were never considered for genotyping. The 58 drug resistant patients had a viral load >1000 copies/ml. The 135 virologically suppressed patients had a viral load <1000 copies/ml.

### Category of patients

Two groups of HIV patients were classified based on viral load. One drug-resistant group (>1000 copies/ml), (N=58), and other virologically suppressed group (<1000 copies /ml to <40 copies/ml to target not detected level), (N=135). A total of 193 patients was included in the sociodemographic and clinical profile analyses.

### Statistical analysis

Patients with drug-resistant (>1000copie/ml) and virologically suppressed patient data, including socio-demographic profiles, socioeconomic profiles, clinical profiles, risk factors, and viral load were recorded on the patient information leaflet and entered into an MS Excel 2010 (v14.0). Descriptive statistical analysis and testing of hypothesis was done using the Chi-square test (P Value) at the 5% level of significance using the statistical software StataSEv11.0 (Stata Corp, College Station, Texas, USA).

### Results

A total of 193 patients was observed in the study whose CD4 counts oscillated around <350 after each six months treatment with first-line anti-retroviral therapy. A detailed description of the sociodemographic, risk factors and clinical profiles of drug resistant and virologically suppressed groups are described in the supplementary Table-1 and supplementary Table-2.

## Common features of drug-resistant and virologically suppressed HIV-1 infected patients

### The entry point of diagnosis

All the drug resistant and virologic suppress had CD4 counts below <350 cubic/mm and were treated with multiple first-line antiretroviral therapy during the attendance of ART center.

### Gender and Age

The highest number of male patients was observed in both groups of patients with an average age of 35 years. No significant difference was observed in the number of patients in the two groups (P=0.094).

### Weight

The average weight of patients in the drug resistant group was 52 years and the virologically suppressed was 50 years.

### Mode of transmission

The heterosexual mode of transmission was highest observed in drug-resistant (38,65.52%) and the virologically suppressed groups (93, 68.89%) respectively.

### Marital Status

The highest number of married couples was observed in drug-resistant (44, 75.86%) and virologically suppressed (101,74.81%) patients compared to other types of marital status.

### Education

The highest number of illiterates was observed in drug-resistant (25, 43.10%) and the virologically suppressed (54,40%) patients.

### Habit of alcohol, smoking, and tobacco

Most of the patients did not use alcohol, smoking, and tobacco in drug-resistant and virologically suppressed groups. In tobacco habit, a significant difference was observed (P=0.026) between the two groups.

### Job and Income

The number of private servants was higher in both groups of patients with an average income of Rs 4000. No significant difference was observed in the monthly income of both drug-resistant and virologically suppressed patients.

### HBV and HCV

The highest numbers of negative patients with HBV and HCV were observed.

### Opportunistic infections

Tuberculosis cases were observed in the highest numbers in the two groups.

### WHO stage

The highest number of patients was at the WHO T1-stage.

### First-line ART

Tenofovir + Lamivudine + Efavirenz (TLE) was administered more times in the two groups of patients.

### CD4 count

The average CD4 was 228.71cubic/mm in drug resistant patients and 240.25 cubic/mm in the virologically suppressed patients. No significant difference {Pr (T>t) 0.65} in CD4 counts was observed between the two groups of patients.

### Adherence

The average adherence to drug-resistant and virologically suppressed patients were 97.36 and 98.34. The detailed description of these features is given in the supplementary table-1 and table-2.

## Discussion

The sociodemographic and clinical data of HIV patients contributed to the sketch of life in the ART Center, Sarojini Naidu Medical College, Agra, North India.

Gender differences in HIV disease progression, the assessment of clinical, social, demographic, virologic, and immunologic factors is very important during treatment of first-line ART [12]. We assessed the gender differences between male and female groups on HIV disease progression over seven years in first-line ART. It is not very clear that how the socio-demographic and clinical profile picture influences the quality of life after confirmation of viral load. Marital status is a major risk of HIV infection and a risk factor for mortality in AIDS. Men were mainly associated with AIDS mortality than women [13]. Among the drug-resistant and virologically suppressed groups, males were observed as the highest number of married couples among the unmarried, divorce, and widow groups. HIV-1 infection is the cause of premature aging between male and female groups. The effect of age on CD4 count and viral load among younger and older patients has been reported. The age mixing patterns and individual levels in age variation provide insight into the persistence and magnitude of HIV transmission dynamics in a field study population [14].

In our analysis, the median age groups of drug-resistant and virologically suppressed, the male age group was the same as the female age. Thus, both the male and female age groups equally affected the HIV transmission in Agra, North India. The CD4 and viral load between male and female groups have no significant difference after over 7 years of first-line ART. Age and sex are important structures in an HIV epidemic study [15]. Therefore, we were well structured the questionnaire data on the basis of age and sex of drug resistant and virologic suppress groups.

Mekuria et al, 2016 [16] reported, the virologic suppression levels can be high without the availability of routine plasma viral load monitoring. Efforts to improve the early detection of HIV positive status and highly active antiretroviral therapy (HAART) initiation for illiterates and those younger patients in a severely immunocompromised state in the two groups. From our study, we compromise the progressive changes in the virologic suppression in long-term first-line ART patients in the Agra region, North India.

In an observational cohort study in the UK, the rates of viral rebound on first-line ART become extremely low over their lifetime [17]. In our report, 67.83% (135/199) patients were on viral rebound that could suppress the virus over 7 years of first-line ART in the Agra region, North India.

In India, the HIV prevalence rate is high among female sex workers and men attending clinics. The HIV-1 epidemic in India is due to the heterosexual mode of transmission [18]. In this case, the heterosexual mode of transmission was a risk factor for HIV transmission in the two groups. This flow of the heterosexual mode of transmission persists in North India. The multi-morbidity was higher in the HIV-positive patients in Odisha, the eastern region of India [19]. In the case of the Agra region, Northern India, the highest incidence of tuberculosis in HIV-positive patients were observed in the two groups. Better adherence with suitable regimen changes during the time of ART leads to a decrease in viral copies among male and female groups. Here, the first-line ART played an important role in the diagnosis to reduce the viral copy number among the 1 to over 7 years of ART. The multiple administration of TLE (Tenfovir +Lamivudine +Efavirenz) suppressed the viral load in 135 patients, out of 199 patients in the ART center of the Sarojini Naidu Medical College, Agra. The TLE regimen effectively suppressed the viral load copies that were taking first-line ART. The spread of HIV in many countries is the most significant impact on education. School-based education is the best practice in HIV prevention for adults and adolescents in low- and middle-income countries, which determines the efficacy of changing HIV-related knowledge and risk behaviors [20]. According to the World Bank group, HIV epidemics are the most devastating impacts on education. HIV/AIDS deprives the strength of education, gradually destroys education quality, drying up countries for skilled workers, and changing the sector cost demand [21]. Education plays an important role in the growth and development of individuals. Education is an important factor that saves life. The education sector played a vital role in ensuring the young’s, orphans, and children’s vulnerability to HIV infection. The education sector is the biggest employer in the world, and discontinuation in education is a big issue in the face of HIV. Education plays an important role in HIV prevention, social support, and protection in the spread of HIV [22]. Education is effective in the male and female groups of HIV patients, creating unemployment opportunities in various job sectors due to mental and psychological stress. The comparison of the education profile of drug resistant and low viral load copies to target not detected levels of patients indicated the highest numbers of illiterates among all educated groups. As the illiterate number was high, the incidence of spread of HIV may be rising in this ART center, Sarojini Naidu Medical College, North India.

The design of this educational sketch in HIV patients is helpful in the spread of HIV in the Agra region, North India. Alcohol use accelerates HIV disease progression was identified in a thirty-month longitudinal study. Alcohol affects CD4^+^ cell count, viral load, and adherence to antiretroviral therapy [23]. Alcohol consumption also influences the toxicity of antiretroviral therapy, adherence, and survival of HIV-infected people [24]. A study reported that alcohol affects ART adherence, quality of life, depression, and external stigma among persons living with HIV in urban India [25]. Agra is an urban city with a higher number of non-alcohol users were observed in this study. Tobacco smoking utilizes a greater impact on HIV-infected patients than uninfected patients. The components of tobacco smoke induce complex pathological changes in different pathways affecting various organ systems in HIV-infected patients [26]. Tobacco smoking is associated with high-risk cancer and cardiovascular problems. It’s also un-suppresses the viral load and CD4 counts in HIV-infected patients [27]. Nonsmokers were higher in both drug-resistant and the virologically suppressed patients in our study. Tobacco use a modifiable risk factor for cancer and cardiovascular diseases in HIV-infected patients. The focus of care to reduce tobacco chewing in HIV-treated patients in a resource-limited setting [28]. Tobacco chewing cases were observed in the drug-resistant and virologically suppressed groups in this ART center, Agra, North India. The use of tobacco and antiretroviral therapy causes several serum biomarkers leading to systemic immune activation in HIV-infected patients [29]. Serum analysis of drug-resistant and virologically suppressed patients will be required in this study.

Thus, the socio-demographic, clinical, and risk factors of drug-resistant and virologically suppressed HIV patients in the Agra region, North India, presents the HIV scenario and will provide a valuable information in the HIV surveillance program.

## Conclusion

The sociodemographic and clinical profile of HIV-1 patients in the ART center, Sarojini Naidu Medical College, Agra, North India could accelerate the care of patients by the physician. Understanding the relationship between the risk factors of drug-resistant and virologically suppressed patients will be helpful in AIDS monitoring and disease progression or death.

## Data Availability

with the article.

https://www.medrixv.org

## Acknowledgement

Indian Council of Medical Research, Govt. of India for providing ICMR-SRFship to Mr. Sushanta Kumar Barik is acknowledged.

