## supplemental table1 for "A study of sociodemographic and clinical profiles of HIV-1 infected North Indian patients"

Supplementary table- 1: Socio-demographic, Socioeconomic, risk factors of drug-resistant (>1000copies/mL) and virologically-suppressed (<1000 copies/mL to target not detected level) patients

| Total number of patients=193  Sex and  Sl. no. of factors |  | Drug resistant patients (>1000copies/mL)  Number of Patients, N=58 | | | | Virologically suppressed patients  ( <1000 copies/mL to Target not detected levels  Number of Patients, N=135 | | | | |
| --- | --- | --- | --- | --- | --- | --- | --- | --- | --- | --- |
|  |  | Number of patients | Percentage (%) | Median | Interquartile range | Number of patients | Percentage  (%) | Median | Interquartile range, | P Value |
| Male | 42 | | 72.41 |  |  | 112 | 82.96 |  |  | 0.094 |
| Female | 16 | | 27.58 |  |  | 23 | 39.65 |  |  |  |
| Age |  | |  | 35 | 12 |  |  | 35 | 15 |  |
| Weight |  | |  | 52 14 | |  |  | 50 | 9 |  |
| Risk Factors:   1. Blood transfusion | 5 | | 8.62 |  | | 5 | 3.70 |  |  | 0.828 |
| 2. Heterosexual | 40 | | 68.96 |  |  | 98 | 72.59 |  |  |  |
| 3. Homosexual | 2 | | 3.45 |  |  | 3 | 2.22 |  |  |  |
| 4. Mother to Child transmission | 1 | | 1.72 |  |  | 1 | 0.74 |  |  |  |
| 5. Probable unsafe injection | 2 | | 3.45 |  |  | 6 | 4.44 |  |  |  |
| 6. Unknown | 8 | | 13.79 |  |  | 22 | 16.30 |  |  |  |
| Marital status:  1.Divorce | 0 | | 0 |  | | 3 | 2.22 |  |  | 0597 |
| 2. Married | 44 | | 75.86 |  |  | 101 | 74.81 |  |  |  |
| 3.Unmarried | 8 | | 13.79 |  |  | 14 | 10.37 |  |  |  |
| 4. Widow | 6 | | 10.34 |  |  | 17 | 12.59 |  |  |  |
| Education:  1.Graduate | 5 | | 8.62 |  | | 9 | 6.67 |  |  | 0.425 |
| 2.Illeterate | 25 | | 43.10 |  |  | 54 | 40.00 |  |  |  |
| 3. Primary School | 14 | | 24.14 |  |  | 48 | 35.56 |  |  |  |
| 4. Secondary School | 14 | | 14.14 |  |  | 24 | 17.78 |  |  |  |
| Habit of Alcohol:  1.Habitual | 2 | | 3.45 |  | | 7 | 5.19 |  |  | 0.697 |
| 2.Never | 40 | | 68.97 |  |  | 81 | 60.00 |  |  |  |
| 3.Past | 8 | | 13.79 |  |  | 24 | 17.78 |  |  |  |
| 4.Social | 8 | | 13.79 |  |  | 23 | 17.04 |  |  |  |
| Habit of Smoking:  1.Current | 6 | | 10.34 |  | | 24 | 17.78 |  |  | 0.106 |
| 2.Never | 44 | | 75.86 |  |  | 81 | 60.00 |  |  |  |
| 3.Past | 8 | | 13.79 |  |  | 30 | 22.22 |  |  |  |
| Habit of Tobacco:  1.Current | 4 | | 6.90 |  | | 22 | 16.30 |  |  | 0.026 |
| 2.Never | 50 | | 86.21 |  |  | 94 | 69.63 |  |  |  |
| 3.Past | 3 | | 5.17 |  |  | 19 | 14.07 |  |  |  |
| 4.Social | 1 | | 1.72 |  |  | 0 | 0.00 |  |  |  |
| Habit of Tobacco:  1. Current | 4 | | 6.9 |  |  | 22 | 16.30 |  |  | 0.02 |
| 2.Never | 50 | | 86.21 |  |  | 94 | 69.93 |  |  |  |
| 3.Past | 3 | | 5.17 |  |  | 19 | 14.07 |  |  |  |
| 4.Social | 1 | | 1.72 |  |  | 0 | 0.00 |  |  |  |
| Job:1. Agriculture cultivator | 1 | | 1.72 |  | | 7 | 5.19 |  |  | 0.260 |
| 2.Agriculture labour | 2 | | 3.45 |  |  | 9 | 6.67 |  |  |  |
| 3.Business | 0 | | 0.00 |  |  | 1 | 0.74 |  |  |  |
| 4.Driver | 0 | | 0.00 |  |  | 1 | 0.74 |  |  |  |
| 5.Govt.servant | 0 | | 0.00 |  |  | 1 | 0.74 |  |  |  |
| 6.House wife | 13 | | 22.41 |  |  | 21 | 15.56 |  |  |  |
| 7.Labour | 1 | | 1.72 |  |  | 9 | 6.67 |  |  |  |
| 8.Local transport worker | 1 | | 1.72 |  |  | 3 | 2.22 |  |  |  |
| 9.Mechanic | 0 | | 0.00 |  |  | 1 | 0.74 |  |  |  |
| 10.No occupation | 12 | | 35.29 |  |  | 22 | 16.30 |  |  |  |
| 11.Petty business | 2 | | 3.44 |  |  | 3 | 2.22 |  |  |  |
| 12.Retd. govt. servant | 0 | | 0.00 |  |  | 1 | 0.74 |  |  |  |
| 13.Self employed | 2 | | 3.45 |  |  | 0 | 0.00 |  |  |  |
| 14.Semiskilled | 0 | | 0.00 |  |  | 3 | 2.22 |  |  |  |
| 15.Service, Private | 19 | | 32.76 |  |  | 38 | 28.15 |  |  |  |
| 16.Skilled worker | 1 | | 1.72 |  |  | 9 | 6.67 |  |  |  |
| 17.Student | 1 | | 1.72 |  |  | 1 | 0.74 |  |  |  |
| 18.Truck driver | 1 | | 1.72 |  |  | 5 | 3.70 |  |  |  |
| 19.Local transport worker | 1 | | 1.72 |  |  | 0 | 0.00 |  |  |  |
| 20.Self employed | 1 | | 1.72 |  |  | 0 | 0.00 |  |  |  |
| Monthly income: |  | |  | 4000 | 5500 | 0 | 0.00 | 4000 | 3000,  0.36 |  |

In the last column P value significance are based on Chi-square analysis
