## Supplemental table2 for "A study of sociodemographic and clinical profiles of HIV-1 infected North Indian patients"

Supplementary table:2

Table 2: Clinical profile of drug-resistant (>1000 copies/mL) and virologically suppressed patients (<1000 copies/mL) to target not detected levels (<40 copies/mL) patients

| Total number of patients=193  Sex and  Sl.no. of factors |  | Drug resistant patients (>1000copies/mL)  Number of Patients, N=58 | | | | Virologically suppressed patients  ( <1000 to Target not detected level (<40 copies/mL)  Number of Patients, N=135 | | | | |
| --- | --- | --- | --- | --- | --- | --- | --- | --- | --- | --- |
|  |  | Number of patients | Percentage (%) | Median | Interquartile range | Number of patients | Percentage  (%) | Median | Interquartile range, | P  Value |
| Opportunistic infections:  1.Hepatitis B Virus (HBV)  A. Negative | 48 | | 82.76 |  |  | 92 | 68.15 |  |  | 0.104 |
| B. Positive | 0 | | 0.00 |  |  | 1 | 0.74 |  |  |  |
| c. Unknown | 10 | | 17.24 |  |  | 42 | 31.11 |  |  |  |
| 2. Hepatitis C Virus (HCV)  A. Negative | 47 | | 81.03 |  | | 93 | 68.89 |  |  | 0.049 |
| B. Positive | 1 | | 1.72 |  |  | 0 | 0.00 |  |  |  |
| C. Unknown | 10 | | 17.24 |  |  | 42 | 31.11 |  |  |  |
| 3. Tuberculosis | 16 | | 27.58 |  |  | 36 | 26.66 |  |  |  |
| 4.Oral Ulcer | 1 | | 1.72 |  |  | 1 | 0.74 |  |  |  |
| 5.Fever | 0 | | 0.00 |  |  | 1 | 0.74 |  |  |  |
| 6. Herpes simplex virus |  | | 0.00 |  |  | 1 | 0.74 |  |  |  |
| 7.Candidasis | 1 | | 0.01 |  | | 1 | 0.74 |  |  |  |
| 8.Tuberculosis and Candi Septimia | 0 | | 0.00 |  |  | 1 | 0.74 |  |  |  |
| WHO Stages:  T1 | 57 | | 98.27 |  |  | 132 | 97.77 |  |  |  |
| T2 | 15 | | 25.86 |  |  | 31 | 22.96 |  |  |  |
| T3 | 23 | | 39.65 |  | | 42 | 31.11 |  |  |  |
| T4 | 19 | | 32.75 |  |  | 28 | 20.74 |  |  |  |
| First line Regimens: |  | |  |  |  |  |  |  |  |  |
| ZLN | 30 | | 51.72 |  |  | 70 | 51.85 |  |  |  |
| ZLE | 14 | | 24.13 |  | | 23 | 17.03 |  |  |  |
| TLN | 29 | | 50 |  |  | 56 | 41.48 |  |  |  |
| TLE | 44 | | 75.86 |  |  | 103 | 76.29 |  |  |  |
| SLE | 6 | | 10.34 |  |  | 13 | 9.62 |  |  |  |
| SLN | 13 | | 22.41 |  | | 18 | 13.33 |  |  |  |
| CD4 count: |  | |  | 228.71 | |  |  |  | 240.25 | 0.65* |
| Viral load (Copies/mL)  1000-  1058687 | 58 | | 100 |  |  |  |  |  |  | 0.99* |
| >40-1000 |  | |  |  | | 18 | 13.33 |  |  |  |
| <40 |  | |  |  | | 29 | 21.48 |  |  |  |
| Target not detected |  | |  |  | | 88 | 65.18 |  |  |  |
| Adherence: |  | |  | 97.36 | |  |  | 98.34 |  |  |

P value given in the last column of the table are based on Chi square analysis. * Two sample t-test
